# Can Social Media Data Be Utilized to Enhance Early Warning: Retrospective Analysis of the U.S. Covid-19 Pandemic

**DOI:** 10.1101/2021.04.11.21255285

**Authors:** Lingyao Li, Lei Gao, Jiayan Zhou, Zihui Ma, David F. Choy, Molly A. Hall

## Abstract

The U.S. needs early warning systems to help it contain the spread of infectious diseases. Conventional early warning systems use lab-test results or dynamic records to signal early warning signs. New early warning systems can supplement these data with indicators of public awareness like news articles and search queries. This study aims to explore the potential of utilizing social media data to enhance early warning of the COVID-19 outbreak. To demonstrate the feasibility, this study conducts a retrospective analysis and investigates more than 14 million related Twitter postings in the date range from January 20 to March 10, 2020. With the aid of natural language processing tools and machine learning classifiers, this study classifies each of these tweets into either a signal or a non-signal. In this study, a “signal” tweet implies that the user recognized the COVID-19 outbreak risk in the U.S. This study then proposes a parameter “signal ratio” to signal warning signs of the COVID-19 pandemic over periods. Results reveal that social media data and the signal ratio can detect the hazards ahead of the COVID-19 outbreak. This claim has been validated with a leading time of 16 days through the comparison to other referenced methods based on Google trends or media news.

## I. INTRODUCTION

The outbreak of an infectious disease threatens society, especially when it evolves into a pandemic in a short period [1]. Coronavirus Disease 2019 (COVID-19), an emerging infectious disease discovered in December 2019 in Wuhan, China [2], has since spread worldwide and engendered an ongoing pandemic [3]. The COVID-19 crisis is particularly severe across the U.S. states and territories, and more than 26 million cases have been confirmed as of February, 2021 [4]. Subsequent news and reports show that the initial U.S. response to the pandemic was slow in terms of preparing the healthcare system, stopping other travels, and implementing testing and measures [5]–[7]. Even in late February, 2020, a Press Briefings from the White House emphasized that the risks to the U.S. public remained low [8]. Due to the delayed response and lack of preparation, the U.S. missed the golden opportunity to contain the COVID-19 outbreak [9]. Early warning systems are critical for controlling this one-in-a-lifetime pandemic.

Early warning for infectious diseases involves analyzing surveillance data with specialized techniques to detect aberrations in the early stages [10]. Surveillance data are the cornerstone of disease warning programs and often originate from laboratory specimens, surveys, dynamic records, sentinel surveillance, and administrative data [10]. Access to these data usually relies on third-party organizations or institutions to initiate provision and sometimes can be costly and time-consuming [11]. New early warning systems can supplement these data with indicators of public awareness like news articles and search queries. The importance of public awareness has been recognized in aiding decision-making across a broad range of research topics, such as environmental conservation [12], health campaigns [13], and emergency preparedness [14]. Analysis of the situational awareness through public opinions can open up new insights to enhance early warning.

Social media platforms are becoming essential tools for decision-makers to communicate information to the stakeholders and harness public opinions from online users. The properties of social media enable individuals to spread information and knowledge and thus make it possible to evoke public awareness on a large scale within a short period [15]. A typical example regarding this COVID-19 crisis is that, before the COVID-19 outbreak, a Chinese ophthalmologist shared the information of a suspected severe respiratory syndrome on social media, and his message raised early awareness about the severity of COVID-19 infections [16]. Understanding such information and opinions created in a particular time and location and its dissemination on social media can provide invaluable resources to feed into early warning systems.

Researchers have demonstrated the potential of aggregating opinions from social media to enhance early warning in many scenarios, such as flood [17], tsunami [18], and pandemic expansion [19]. Building on the existing body of knowledge related to the utility of social media in aiding early warning, this study conducts a retrospective analysis and explores the potential of leveraging public awareness through Twitter data to signal early warning for the U.S. COVID-19 pandemic. While this approach does not establish a response mechanism to cope with disease control and prevention, it provides public and private sectors a timely and efficient instrument to detect the hazards by “crowdsourcing” public opinions, thereby enabling them to take early preparedness to contain the pandemic.

## II. LITERATURE REVIEW

### A. SOCIAL MEDIA IN EMERGENT EVENTS

Social media data are often obtained from Twitter, Facebook, Instagram, and other web or mobile platforms. Luna and Pennock summarized three crucial benefits that social media bring in emergency management: increasing situational awareness, accelerating information diffusion, and monitoring activity and status [15]. Owing to these benefits, researchers have demonstrated the potential of utilizing social media data to assist the management of emergent events [20]. Multiple studies evaluate tweet volume with the target domain words to monitor natural hazard events (e.g., flood, earthquake, and hurricane) [21], [22]. A typical application is to detect worldwide earthquakes through monitoring the intensity of earthquake-related messages sent on Twitter [22]. With the advancement of natural language processing and machine learning techniques, recent studies have combined them with social media data to aid the analysis. For example, Chao *et al*. proposed a hybrid machine learning pipeline with named entity recognition that leverages relevant tweets to uncover the evolution of disaster events across different locations [23]. Bhoi *et al*. proposed a prototype system that integrates word embedding with deep learning models to analyze emergency-related tweets to enhance resource management [24].

In the field of infectious disease, one primary area focuses on emergency communication and cooperation. Perhaps this is because social media provide timely channels for government agencies to communicate information to stakeholders and for individuals to share such information with families and friends. This reveals importance in the pandemic outbreaks. Yang and Sun investigated the health policy under COVID-19 and discussed how the role of public voice on social media can help the government to promote policy evolution [25]. Abrams and Greenhawt manifested that one possible way to ensure appropriate risk communication amid the COVID-19 pandemic is using social media channels to provide an ongoing and consistent media presence [26]. Kim and Hawkins suggested that social media strengthen shared awareness and contribute to positive health prevention behaviors amid the 2019 U.S. measles outbreak. In particular, social media expression and reception can help promote preventive hygiene intention [27].

### B. SOCIAL MEDIA IN EARLY WARNING AND DISEASE SURVEILLANCE

The extensive applications of social media unravel new insights into the early warning systems in many scenarios, like flood [17], tsunami [18], and pandemic expansion [19]. Understanding where the information has been posted and analyzing related contents created in a particular time window can provide a valuable resource to feed into early warning systems [28]. Multiple efforts capitalize on the analysis of textual content, information dissemination, or sentiment patterns to signal early warning for an emergent event [29]– [31]. A typical example tracked the sentiment changes based on users’ geolocation information, aiming to facilitate a more robust early warning system for hurricane disasters [29]. Similar to this study, Kitazawa and Hale examined millions of tweets during a typhoon crisis occurred in Japan. They found that emergency warnings were likely to have people be attentive, and the expected shift of public attention on social media could serve as indicators for such alarm [32].

Early warning for infectious diseases involves analyzing surveillance data in the early stages of a pandemic [10]. Studies using social media data for surveillance of disease outbreaks are closer to this present work. One popular approach is to monitor online health-seeking behaviors, such as search queries to web search engines [33]–[35], or to detect aberrational trends via the volume of related online news [36]. For example, Ginsberg *et al*. specifically discussed how to use Google search queries to track influenza-like illness amid the epidemic [35]. Other researchers have applied a similar method to detect epidemics or pandemics through monitoring search queries [33], [34]. A recent article validated the credibility of using internet searches and social media data to predict COVID-19 outbreak by displaying a strong correlation between search indexes with subsequent reported COVID-19 cases [37].

The other common approach leverages textual analysis techniques to handle social media data for disease surveillance [38]–[40]. The objective of these studies is to establish a real-time surveillance system or an event detection system to track the infections [41], [42]. The importance of this application is evident, and thus the frequency of these applications is obvious. For example, Aramaki *et al*. extracted millions of influenza-related tweets and applied machine learning models to classify the tweets into either symptom-related or -unrelated [38]. Their experiment results displayed a high correlation with influenza cases that outperforms the Google search method. Rosa *et al*. adopted topic modeling techniques to analyze users’ behavior changes in online social networks [43]. They have applied their model to detect the COVID-19 pandemic in the early stages. Overall, these achievements demonstrate the credibility of using social media data to predict the hazards of disease outbreak.

### C. RESEARCH GAP AND OBJECTIVE

With the reviewed studies, it is clear that early warnings based on social media data requires the detection of an aberrational pattern, such as a shift of people’s thoughts or behaviors in the early stages. Most of the attempts combining social media data aim to provide real-time surveillance, which mainly relies on the classification of symptom-like messages. However, these models cannot be used to signal early warnings when the symptoms are not widely recognized. While the search engine-based methods have demonstrated the possibility to predict the hazards, they may not be applicable when aberrations are not discernible at the beginning of a crisis. More importantly, with the reviewed works, there has been minimal focus on how to leverage public opinions via social media or how to quantify the awareness level to release warning signals to a pandemic.

To fill this research gap, this study attempts to use social media data obtained from the Twitter platform and explore the potential of harnessing public opinions to promote early warning systems. This study conducts a retrospective analysis through reviewing the early stages of the U.S. COVID-19 pandemic. The remainder of this study is organized as follows. Section 3 presents data preparation procedures and introduces tools and classifiers to build the pipeline to classify each tweet into a “signal” or a “non-signal.” The “signal” tweet implies that the user thought the COVID-19 outbreak was likely to happen in the U.S., while a “non-signal” tweet implies that the user discussed COVID-19 related topics but did not show clear concerns regarding the risks of the outbreak. Section 3 also explains the parameter “signal ratio” and the “tipping point” concept to deliver warning signs. Section 4 exhibits the results from both temporal and spatial dimensions and displays the aberrational patterns and concern shifts. Section 5 discusses the findings, significance, implications, and limitations.

## III. DATA AND METHODS

### A. DATA PREPARATION

Twitter data reflect the information that users choose to share in public. In this study, we utilized the Twitter Standard Search API with the search term “Coronavirus” to download related tweets in the date range from January 20 to March 10, 2020. We selected this date range before the World Health Organization (WHO) announced that COVID-19 was a pandemic on March 11, 2020 [44], aiming to illustrate the feasibility of using the indicators of social media data to deliver early warnings. Since the WHO officially named “Coronavirus” as “COVID-19” on February 11, 2020 [44], we did not use “COVID-19” as the search term concerning the consistency of data scraping. Although there were other terminologies to describe this pandemic, such as “Wuhan virus,” “Chinese virus,” and “pneumonia,” we chose not to use them either as they contained discrimination meanings or were not specific to this disease. Moreover, using the keyword “Coronavirus” could generate enough data representative of public opinions, and thus we selected this keyword to collect tweets.

However, Twitter data, with limitations on contents, are currently and unlimitedly available without fee for one-week following their postings with Twitter Standard Search API. To ensure the consistency of the data associated with the target topic, we used the API to collect related tweets daily from the beginning of the study period. These daily scraped data were stored in simple JavaScript Object Notation (.json) formats and converted to Excel (.xlsx) files for subsequent processing. This scraping process resulted in an original dataset with 83,773,379 records. Since the original data included the tweets containing “Coronavirus” from worldwide, we filtered in records with users’ registration locations only implying a location in the U.S. For example, registration locations like “USA,” “Washington DC,” “NYC,” “California,” and “Los Angeles” all indicate locations in the U.S. This filtering process reduced the dataset to a total of 14,702,253 records with 3,598,708 unique tweets given that retweets are of the same textual content.

Twitter provides four types of tweets that people can post on its platform: general tweets, mentions (i.e., quotes), replies, and retweets [45]. A general tweet is a message posted on Twitter containing text, photos, a GIF, or a video. A mentioned tweet quotes another account’s Twitter username, and a reply is a user’s responding text to another user’s tweet [45]. These three types of tweets contained a user’s original texts and attributions and were considered as original tweets in this study. Unlike these three types of tweets, a retweet is a reposting of another user’s tweet [45]. For the present study, we did not remove retweets, given that a user who reposted another user’s tweet could hold the same opinion or recognize the same risks regarding the COVID-19 outbreak. A tweet with more retweets could also indicate that its described information was recognized by more people on Twitter.

To establish the training dataset, we selected the top 5,000 most frequently retweeted tweets and manually classified each tweet into a “signal” or a “non-signal” class. In this study, a “signal” tweet implies that the user thought the COVID-19 outbreak was likely to happen in the U.S. possibly due to the lack of preparation to contain the transmissibility of the virus. A “non-signal” tweet implies that the user discussed COVID-19 related topics in the tweet but did not show any concern regarding the risks of the outbreak. Based upon our review on the factors to control disease outbreak [46]–[48], subsequent news and reports [5]–[7], and manual inspection for the 5,000 tweets, we summarized three major types of tweets that could be used to signal the risk of COVID-19 outbreak, as exhibited in Table 1. For example, according to the news published in the Associated Press on April 13, it attributed the U.S. COVID-19 outbreak to slow responses, in terms of preparing the healthcare equipment and testing kits, and underestimation of the threat [5]. These “signal” tweets reflect how the public awareness emerged from Twitter can be used as “human-sensors” to signal the early warnings of COVID-19 outbreak.

In addition, we selected the top 5,000 most frequently retweeted tweets rather than a random subset to build the training dataset based upon two considerations. First, these 5,000 tweets are most likely to be reflective of impactful twitter data on the dataset as they received most retweets. More importantly, labeling these 5,000 tweets could ensure more accurate classifications on the whole dataset. It was worth noting that they comprised 5,931,904 (40.3%) of the full dataset (retweets were considered as the same content in this study). In other words, once the trained model got a training accuracy of 99%, retweets of these top 5,000 tweets could be almost correctly classified. Therefore, this treatment could ensure a higher classification accuracy on the whole dataset.

**TABLE 1.** Three major types of tweets that were classified as “signals” for sensing hazards.

| Type | Description | Tweet Example |
| --- | --- | --- |
| 1 | Suggest that the U.S. was not prepared, such as lack of testing kits or equipment. | *Is this a secret? As of a week ago, only 500 people were tested. The lack of testing allowed the coronavirus to spread within the US undetected.* |
| 2 | Hold the opinion that the risk of the pandemic was downplayed, and the real hazard was hidden | *I'm sure some people think there is a lot of talk about Coronavirus. I'm frankly surprised there's not more. Italy sounds completely overwhelmed. That's looking like us I two weeks. The President is continuing to downplay the threat, which will get more people sick.* |
| 3 | Show worries for disease spreading based on daily observation or personal experience. | *I just landed at JFK after reporting on #coronavirus in Milan and Lombardy—the epicenter of Italy's outbreak—for @vicenews. I walked right through US customs. They didn't ask me where in Italy I went or if I came into contact with sick people. They didn't ask me anything.* |

Fig. 1 presents the framework to implement the text classification model on the dataset. We divided the dataset into two domains: 1) those 5,000 human labeled tweets comprised 40.3% of the full dataset and were used to train the classification model, and 2) the rest 59.7% tweets were automatically labeled by the trained classification model. We developed the text classification pipeline following four major steps: text labeling, text augmentation, text vectorization, and classification.

**FIGURE 1.**
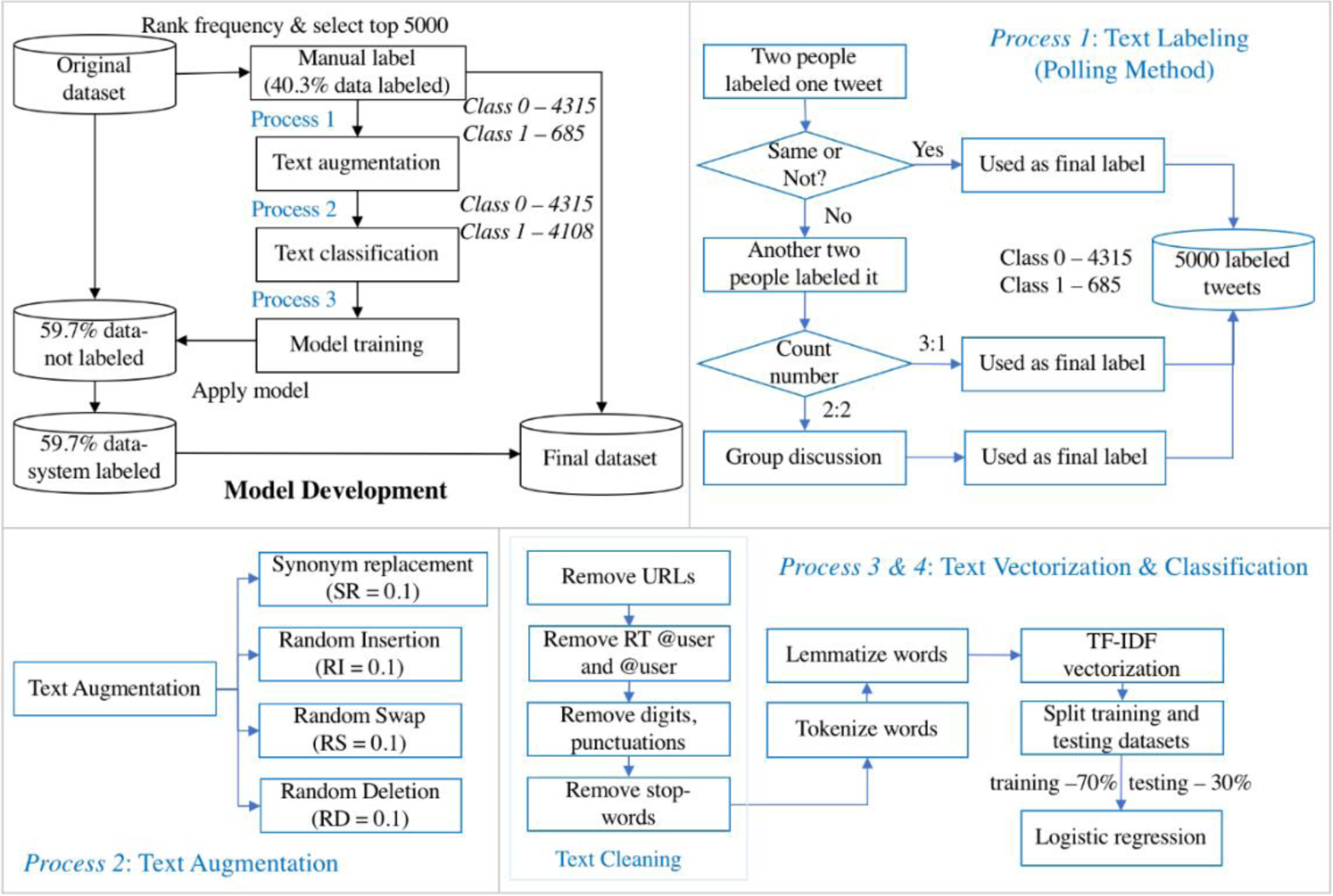
Research framework for the implementation of the classification method.

### B. HUMAN TEXT LABELING

The authors manually reviewed the 5,000 tweets and categorized each tweet into a “signal” (class 1) or a “non-signal” (class 0) through three rounds of labeling (Fig. 1). In the first round, two handlers formed a group, and each group was responsible for labeling half of the 5,000 tweets. The label criteria were based on our reviews on the factors to control disease outbreak and subsequent news and reports, as Table 1 exhibits. When a tweet received the same label from both handlers, it was considered as the final label for this tweet. When there was disagreement on the labels, another two handlers in the research team came to label this same tweet. The class of this re-labeled tweet was determined following the majority of the four views. However, when this tweet again received two same labels for both class 0 and class 1, it was put on the discussion board and determined by all participated handlers. As a result, 4,315 out of 5,000 tweets were classified as “non-signal” (class 0) and 685 tweets as “signal” (class 1), in which 198 tweets were put on the discussion board that were decided by all the handlers.

### C. TEXT AUGMENTATION

Before text augmentation, we applied several steps to clean the tweets, as presented in the box “Text Cleaning” in Fig. 1. First, short URLs, @username, RT @username, digits, emojis, and stop words, punctuations were removed in a tweet. Next, each tweet was tokenized into a list of separate words and characters. Since the words in a tweet were written in different forms, the tokenized words were converted to their stemming forms. This process is known as lemmatization. We employed the Natural Language Toolkit (NLTK) python package to complete this text cleaning process [49].

Among the 5,000 tweets, 13.7% of them (685 out of 5,000) were classified as “signal” tweets, while 86.3% (4,315 out of 5,000) were classified as “non-signal” tweets. Such severe imbalance in the training dataset posed an issue for text classification. That is, the trained a model could produce reduced performance for the minority class. Specifically, the trained model could achieve as high as 85% accuracy even though all outputs were predicted as “non-signal” tweets (class 0). As a consequence, the trained model failed to detect a tweet as a “signal.” There are two practical methods to tackle this issue, namely downsampling and upsampling. Downsampling balances classes by training on a proportionately small subset of the majority class samples, while upsampling balances classes by increasing the size of the minority class samples. Since downsampling might result in a significant loss of information for the model to capture the “non-signal” cases in this research context, we determined to select upsampling as the text augmentation method to ease the imbalance and increase training size.

Among all the upsampling techniques, we adopted a straightforward technique called Easy Data Augmentation (EDA). This technique requires no model to be pre-trained on external datasets and is capable of substantially improving the performance, especially on a training set with less than 500 samples [50]. The EDA model uses four operations to increase the data volume [50], as exhibited in Table 2. We followed the recommendations in their article and set the ratio as 0.1 for each operation. Based on the imbalance ratio ((4,315-685)/685=5.3) in the 5,000 labeled samples, we set the times for augmentation of the minor class to 5. This process generated a training dataset with “signal” and “non-signal” tweets approximately equivalent, including the original labeled 4,385 “non-signal” tweets and the augmented 4,110 “signal” tweets.

**TABLE 2.**
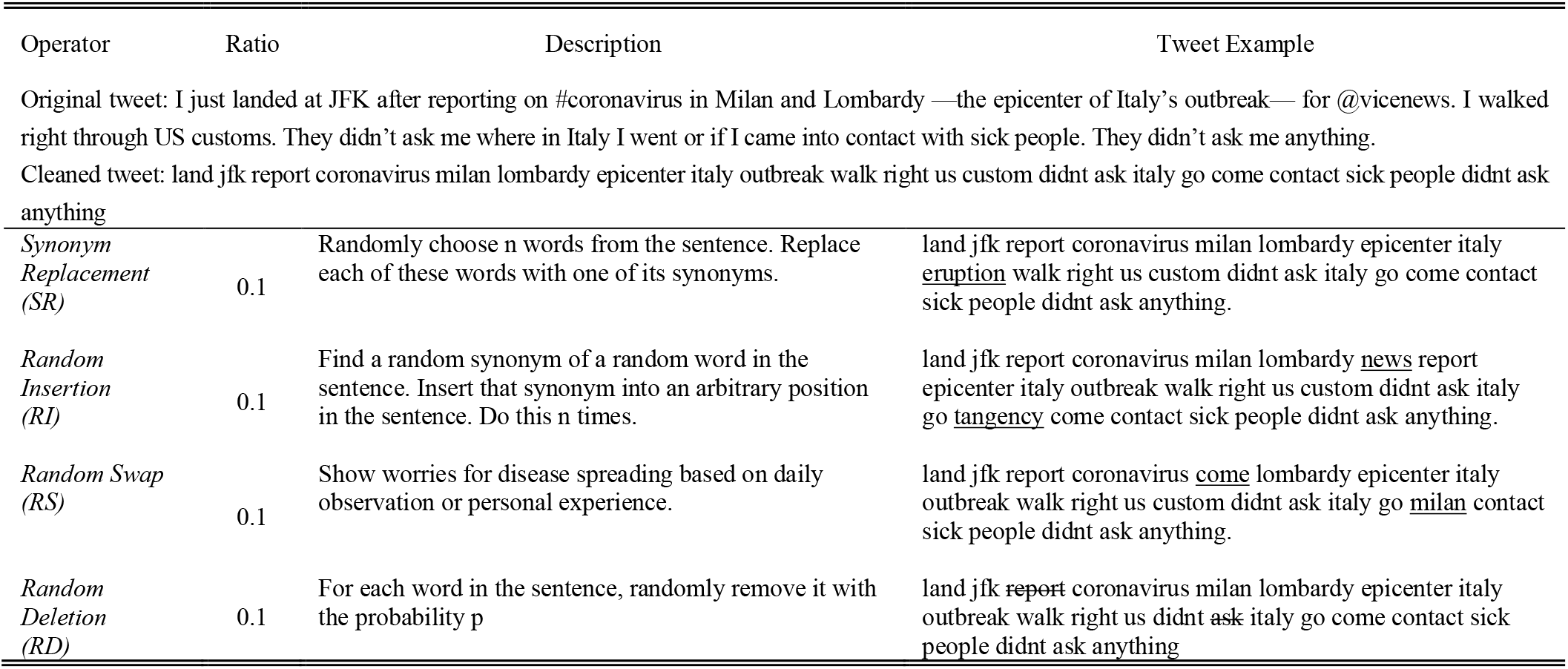
Text augmentation operators and examples.

### D. TEXT VECTORIZATION

Term Frequency-Inverse Document Frequency (TF-IDF) was utilized in this study to convert each tweet into a vector of features. TF-IDF is an efficient and effective term weighting method widely applied in the fields of text similarity, text classification, and information retrieval [51]. TF-IDF defines the importance of key terms as [51],

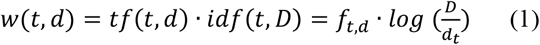

in which, *w*(*t, d*) represents the weight of word *t* in tweet *d*; *f*_*t,d*_ denotes the frequency of word *t* in tweet *d*; *D* is the total number of tweets; and *d*_*t*_ is the number of tweets that word *t* appears. Words with high TF-IDF value imply strong relationships with the tweets where they appear. Although TF-IDF does not capture word positions or semantic similarities in a sentence, it is an efficient and simple algorithm for matching words in a query to documents [52]. Due to its robustness and fast computation, TF-IDF is especially useful when dealing with a large set of Twitter data in different text lengths. In this research context, all words in a tweet were represented with numerical information, and each tweet was mapped into a numerical vector for classification.

### E. TEXT CLASSIFICATION

After each tweet was converted to a vector of features using TF-IDF, we applied several machine learning classifiers provided by Scikit-learn Python library [53], including Random Forest (RF), Logistic Regression (LR), Support Vector Machine (SVM), and Naïve Bayes (NB) to build the pipeline for text classification. Given that LR achieved the highest testing accuracy as shown in Table 3, we specifically explained this algorithm in this section. LR is a well-developed machine learning technique borrowed from statistics, which is intended for binary classification. Logistic regression is characterized by a logistic function to model the conditional probability of the label Y given independent variable X, *P*(*Y*|*X*) [54]. Given a tweet vector, the conditional probability of a tweet classified as class 1 or class 0 can be modeled using the sigmoid function [54]:

**TABLE 3.** Performance measurement of different models.

| Type | RF +<br>TF-IDF | LR +<br>TF-IDF | SVM +<br>TF-IDF | NB +<br>TF-IDF |
| --- | --- | --- | --- | --- |
| *Precision* |  |  |  |  |
| Class 1 | 0.89 | 0.89 | 0.87 | 0.81 |
| Class 0 | 0.87 | 0.98 | 0.86 | 0.97 |
| *Recall* |  |  |  |  |
| Class 1 | 0.86 | 0.98 | 0.86 | 0.97 |
| Class 0 | 0.90 | 0.87 | 0.87 | 0.78 |
| *F1-score* |  |  |  |  |
| Class 1 | 0.88 | 0.93 | 0.86 | 0.89 |
| Class 0 | 0.88 | 0.92 | 0.87 | 0.86 |
| *Training* | 0.93 | 0.99 | 0.90 | 0.93 |
| *Testing* | 0.88 | 0.93 | 0.86 | 0.88 |

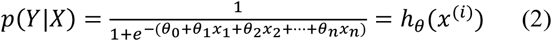

where *Y ∈* {0,1}, *Y* = 0 denotes “non-signal” (class 0) and *Y* = 1 denotes “signal” (class 1); *x*^(*i*)^ = (*x*_1_, *x*_2_, …, *x*_*n*_) denotes the converted vector of the i^th^ tweets using TF-IDF method; *θ* = (*θ*_0_, *θ*_1_, *θ*_2_, … *θ*_*n*_) denotes the model parameters that need to be trained. The sigmoid function is the core of logistic regression that maps the vectors of tweets to one of the binary classes. The loss function *J*(*θ*) applied in this study is cross-entropy loss, and its mathematic formula is presented below.

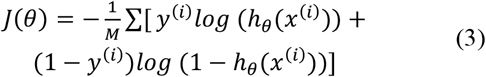

where *h*_*θ*_(*x*^(*i*)^) is the probability value computed by sigmoid function, *i* denotes the i^th^ data point (i.e., tweet) in the dataset, *θ* is the model parameters, and *M* is the total number of samples. This cross-entropy loss increases when the predicted probability deviates from the actual labels. This built pipeline classifies a tweet with the value of sigmoid function > 0.5 implying *Y* = 1 (“signal”) and <0.5 implying *Y* = 0 (“non-signal”). To implement the text classification, we split the training and testing size into 70% and 30% of the augmented dataset. According to Gholamy *et al*., empirical studies show that the best results are often obtained when using 20∼30% data for testing, and the remaining 70∼80% for training [55]. The 70/30 split in this study made the classification pipeline trained with enough data whilst more testing data made the error estimate more accurate.

### F. PERFORMANCE MEASURES

Precision, Recall, and F1-score were applied to assess the classification performance since these indexes could better reflect the performance on unbalanced classes. Precision measures the fraction of true positive cases over the retrieved cases that a model predicts, while recall is the fraction of true positive cases over all the relevant cases that are actually identified. The F1-score is a rating of test accuracy, representing a combination of Recall and Precision [56]. Higher F1-score implies higher testing accuracy [56]. The formulas of Precision, Recall, and F1-score are presented in (4), (5), and (6), respectively. Performance on the testing set of these classification pipelines is exhibited in Table 3. The LR classifier outperformed the other three classifiers and thus was selected for the following investigation, demonstrated by higher F1-scores and testing accuracy. It is apparent that other classifiers could easily have been applied once their performance warranted their choice in other cases.

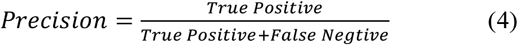

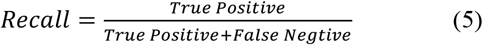

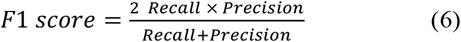

### G. ARNING LEVEL CALIBRATION

In this study, we propose a parameter, “signal ratio” to quantify the public awareness of the COVID-19 outbreak over periods. The signal ratio is measured as a fraction of the number of “signal” tweets divided by the sum of both “signal” and “non-signal” tweets (7). We then adopted the concept of “tipping point” from sociology to signal the warning signs. The concept of “tipping point” refers to the moment when an idea, trend, or social behavior crosses a threshold and spreads like wildfire [57]. The idea of using “tipping point” as the warning signal is broadly applied in monitoring climate change [58] as well as identifying social issues [59], [60]. Previous studies have put this “tipping point” threshold anywhere from 0.10 to 0.40, according to a report published in *Science* [61], which demonstrated that when the committed fraction grows beyond a critical value of 0.10, there is a dramatic decrease in the time for the entire population to adopt the committed opinion [62]. That is to say, when the number of committed opinion holders is about 10 percent of the population, their belief is likely to be adopted by the majority of the society [62]. Although the committed fraction in their study referred to the committed agents in a network (e.g., Twitter users in our study) and was calculated based on modeled networks rather than real networks (e.g., Twitter network), we considered the fraction of “signal tweets” to approximately reflect the fraction of “opinion holders” in the Twitter network. At this point, previous studies have also used the tweet volume to represent the public opinions in terms of political and social issues [63]–[65], and these studies support the feasibility of our assumption. Moreover, we set 0.1 as the critical value to signal the warning based on multiple attempts to explore the results-oriented performance with respect to the warning patterns. Fig. 2 shows a “scattered” warning pattern at the beginning of our study period and a “continuous” warning pattern later, and this evident pattern shift could suggest an aberration for early warning systems.

**FIGURE 2.**
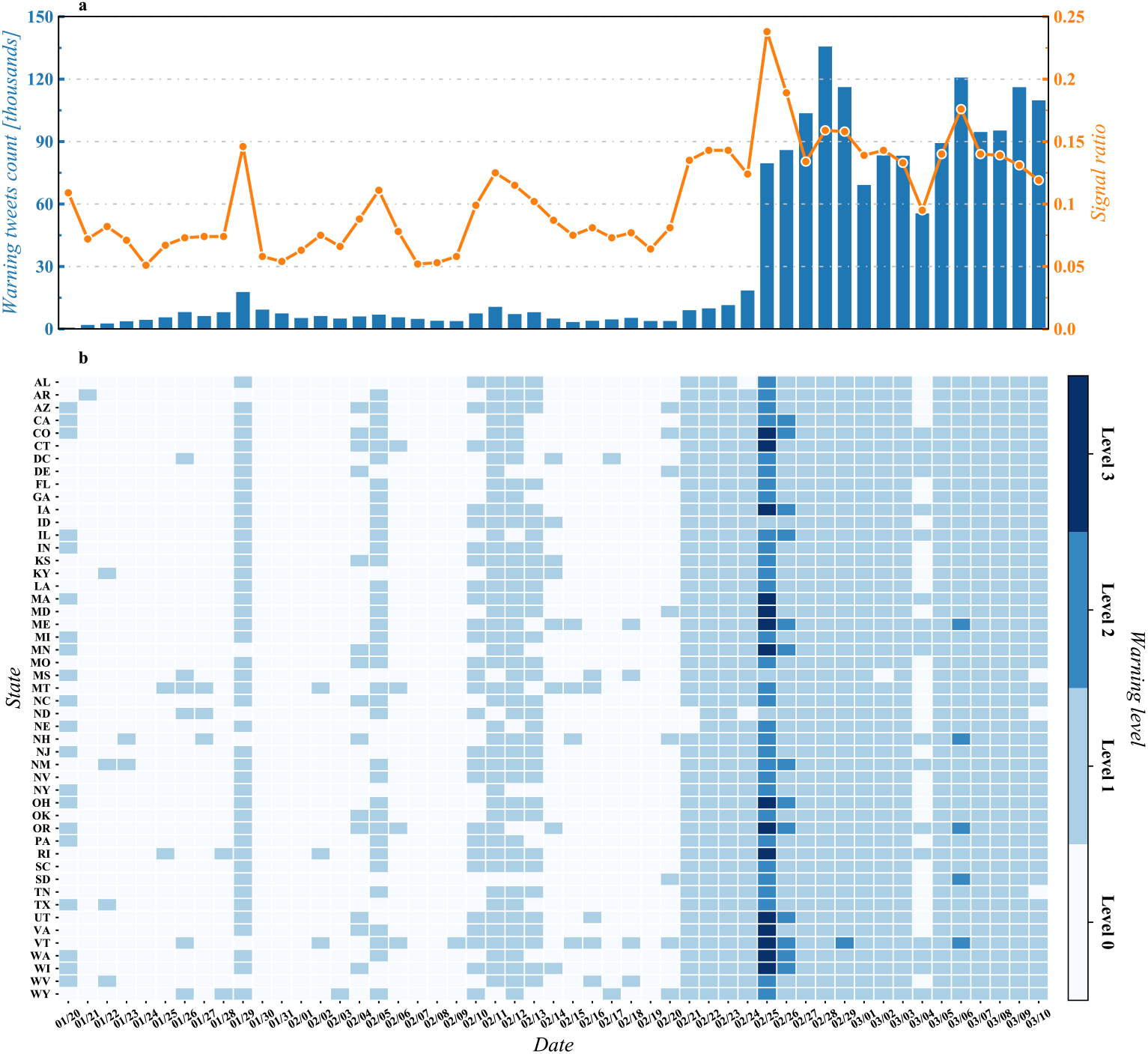
Daily signal ratio and warning level for each state.

For these considerations, this study uses 0.10 as the threshold to signal warning signs in this specific research context. That being said, the signal ratio delivers a warning when “signal” tweets occupy a proportion of 0.10 of all COVID-19 related tweets. To further differentiate the warning levels, we treated a signal ratio > 0.20 as warning level 2 and > 0.25 as warning level 3. This warning calibration can signal warning signs even with low tweet volume in the early stages.

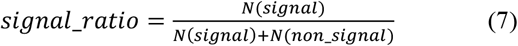

## IV. RESULTS ANALYSIS

### A. TEMPORAL AND SPATIAL RESULTS

We computed the daily signal ratio based on (7) and presented it in Fig. 2a. To calculate the state-level signal ratio, we firstly grouped the data based on the users’ registration locations. For example, tweets with the following registration locations, “California, USA,” “Los Angeles,” “California,” and “Santa Monica CA,” all indicate California State. We then created a heatmap to track warning levels for each state, as shown in Fig. 2b.

At the beginning of the study period, when the Chinese government announced a national-wide emergency, people on Twitter started to sense the risks of COVID-19 transmission. Correspondingly, the signal ratio delivered a few early warning signs even though the tweet volume remained low at this time. The signal ratio reached 0.15 and delivered a second clear warning signal on January 29, and this warning sign was captured by most states, according to Fig. 2b. From February 11 to February 13, the signal ratio crossed 0.10 in three consecutive days. Until late February, only a few cases were officially confirmed in the U.S. that were all imported overseas, and the government emphasized that the risks to the U.S. public remained low at that time [8]. However, these signs that emerged from public awareness, although scattered, provided early warning information that could assist the decision-makers in containing the disease outbreak.

After the first community transmission reported on February 20, the volume of “signal” tweets dramatically increased and spiked on February 28. The risk of COVID-19 outbreaks was extensively recognized in the Twitter community. The signal ratio appeared to show a continuous warning pattern, as presented in Fig. 2. The signal ratio reached the highest level for the entire U.S. on February 25, showing warning level 3 for some states and level 2 for most U.S. states. The Europe’s outbreak in Italy enabled U.S. people to recognize the high risks. However, President Trump remained publicly optimistic about the virus [66], which raised wide concern regarding the U.S. preparation for the outbreak. Although the signal ratio subsided somewhat the next day, its value fluctuated around 0.15 over the following days. One possible explanation was that people were aware that the COVID-19 situation was far worse than previous estimations. This continuous warning pattern reflected public’s constant worries concerning that the U.S. might not be prepared to respond to the COVID-19 outbreak. It also released a danger signal for the decision-makers that the disease was about to leap out of control.

In addition to temporal analysis, we plotted the average state-level signal ratio on geography, as exhibited in Fig. 3. The state-level signal ratio was computed based on the classified tweets with the same state registration information. Overall, the signal ratio ranges from 0.10 to 0.17 across states, among which Vermont (0.17), Oregon (0.16), Minnesota (0.15), Delaware (0.15), Wisconsin (0.15), and Wyoming (0.15) have a higher signal ratio than other U.S. states. Twitter users from states with higher signal ratio were more aware of the COVID-19 risks. According to Fig. 3, states located in the West, Midwest (especially East North Middle), and the Northeast region displayed a higher signal ratio when compared to states located in the South and Middle areas.

**FIGURE 3.**
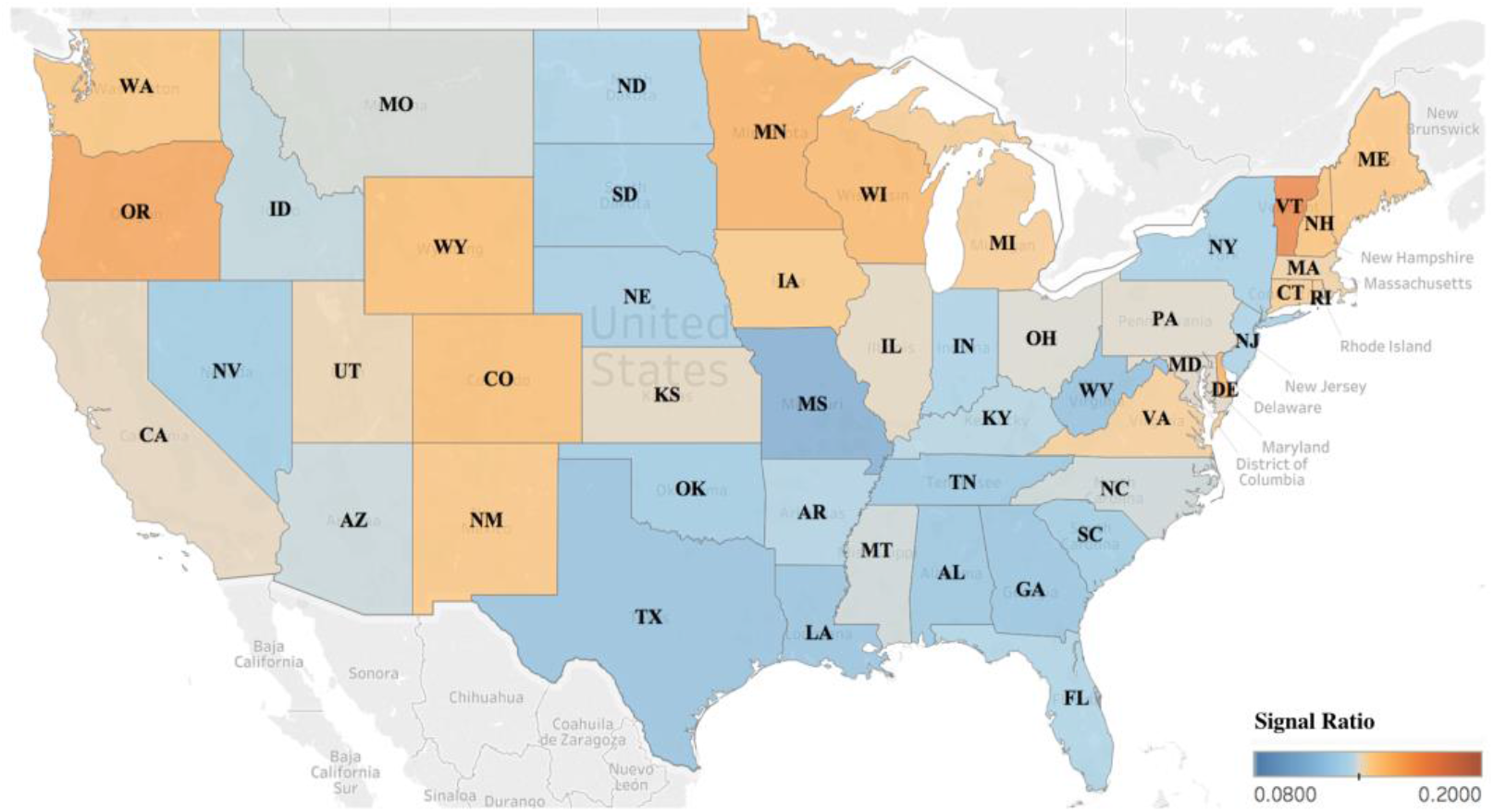
Average state-level signal ratio in the 48 contiguous U.S. states.

### B. DEPICTION OF PUBLIC CONCERNS

In this section, we conducted an exploratory analysis to shed light upon how the public concerned about the COVID-19 outbreak and how the patterns shifted during the study period.

To do so, we manually reviewed the top 20 tweets that were most frequently reposted from each of the “warning” days. After that, we summarized the major concerns and presented a cluster analysis to depict the shifts of public awareness, as shown in Fig. 4.

**FIGURE 4.**
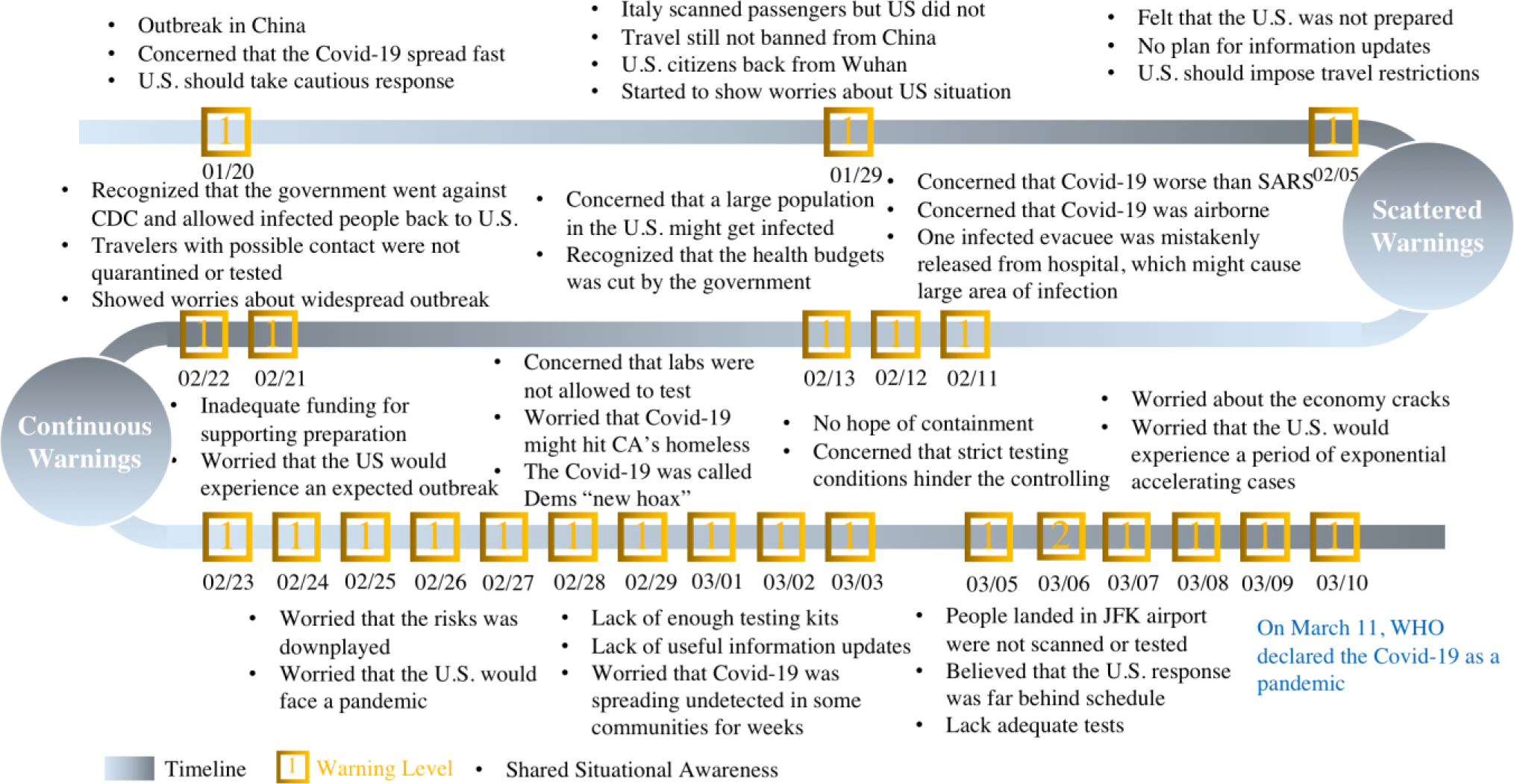
Shared situational awareness emerged from “signal” tweets.

In late January, the COVID-19 outbreak in China raised early concerns about the risk. People on Twitter suggested the government to take precautionary measures until the severity and nature of COVID-19 was understood. Another two events also raised early awareness: 1) Italy scanned passengers, but the U.S. did not, and 2) citizens came back to the U.S. from Wuhan, China, the center of the outbreak. In early February, as more details of COVID-19 were reported, people showed anxiety that the virus was far more contagious and could be extremely harmful to the society. During this period, the signal ratio presented a “scattered” warning pattern. The public awareness focused on the virus mechanism (e.g., viral transmission) and the outbreak in other countries and the U.S. connections with them (e.g., travel restrictions).

Nevertheless, starting from February 20, the signal ratio presented a “continuous” warning pattern. We probed into those “signal” tweets and discovered that the public awareness recognized four major hazardous phenomena: 1) supply shortages, such as lack of testing kits and health equipment; 2) late response and insufficient preparation, such as late information updates and inadequate funding for supporting preparation; 3) downplayed risks and hidden facts; 4) reports of the first case in many states. In early March, many people posted pessimistic messages stating that the U.S. had already lost the opportunity to contain the outbreak. Meanwhile, people were worried about that the U.S. would experience an exponential acceleration of reported cases, and the U.S. economy would crack. This observation reflected that the public awareness emerged on social media might recognize the hazards considerably earlier than the government official statements. Apart from the shift of the warning patterns, the focus of public awareness shifted from the concerns about the COVID-19 outbreak in other countries to the fear of the viral transmission, followed the distrust of government capabilities to deeply pessimistic views of the U.S. situation. It was further noted that such pattern shifted from the focuses on external factors (e.g., outbreaks in other countries and virus mechanism) to the focuses on internal factors (e.g., lack of testing kits and equipment, distrust of government, and the prospect of economy and society).

There are a couple of possible reasons to explain such observations. First, social media gave U.S. citizens visibility to news and opinions concerning COVID-19 from other countries. For example, the COVID-19 outbreak in China raised significant concerns about the situation in the U.S. as people were aware that the virus was contagious and could be easily transmitted among communities. Second, people on social media might project U.S. situations based on their own observations or traveling experience. A typical example was that, a Twitter user expressed her concern about the U.S. preparedness after landing the JFK airport from Italy, since the airports took no anti-epidemic measures (e.g., temperature scanning and social distance) for passengers. As a result, her concern raised an extensive discussion on Twitter regarding the U.S. preparation for the outbreak. Third, many opinions that emerged on social media came from epidemiologists, clinicians, statisticians, survey specialists, and government officials. Their thoughts could provide useful insights to understand the risks of COVID-19 in the early stages. For example, an infectious disease expert, Michael Osterholm, warned in early March that the U.S. was ill-prepared to combat the COVID-19 and would face a pandemic [67]. The properties of social media enabled these opinions to spread quickly through the internet and thus made it possible to evoke public awareness within a short period. These explanations further unravel the rationality of using indicators of public awareness through social media to enhance early warning.

### C. COMPARISON TO OTHER RESOURCES

This section attempts to check the credibility of this proposed approach by examining the signal ratio with other referenced methods. The first referenced method was proposed by [68], with an assumption made that the number of related news posts from media could reflect the warning level to a certain extent. In their study, a news post was counted as long as its title contains at least one word related to COVID-19, such as pneumonia, coronavirus, and SARS. Their data resource comes from the Global Database of Events, Language, and Tone (GDELT), which stores metadata for international news [69]. We followed their approach to retrieve related index information and presented it in Fig. 5a (solid blue line). The second method follows the approach based on the search queries to Google [35]. The data resource was extracted from the Search Volume Index of Google Trends. To visualize these indexes in the same range, we normalized the Index of Google Trends into the range between 0 and 1 and plotted it in Fig. 5a (dotted blue line).

**FIGURE 5.**
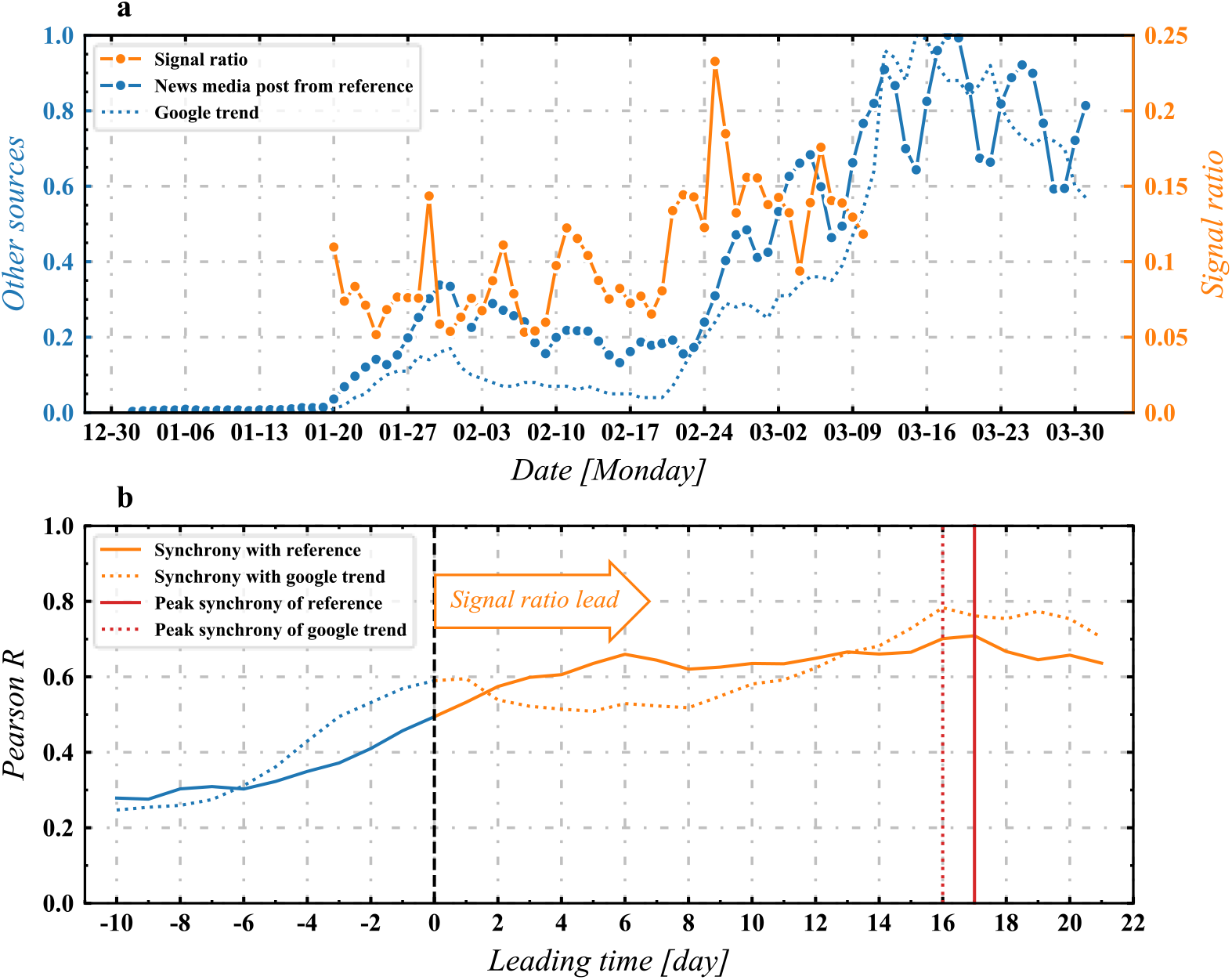
Validation through comparison with the multiple information resources.

Upon initial inspection, the relations between signal ratio and the referenced indexes might not be evident. Therefore, we computed the time-lagged cross-correlations and presented the relations in Fig. 5b. The time-lagged correlation can help quantify the synchrony between two sets of time series data and unfold a clearer view of the connections. In Fig. 5b, the leading time represents how long the signal ratio is ahead of the other two referenced indexes. A higher correlation suggests a higher alignment under a specific lagging date. According to Fig. 5b, the signal ratio displays a moderate to strong correlation with other referenced indexes when it leads the time lag. In particular, the correlation rises to 0.7 when the signal ratio leads 17 days ahead of the referenced method based on media news and 0.8 when the signal ratio leads 16 days ahead of the Google Trends. While the referenced methods based on Google Trends or media news provide quantitative information to support early warnings, their applications may be limited to the situations where the monitored trends (e.g., banner headlines, news subscriptions, and search queries) show clear aberrations. Moreover, the time-lagged cross-correlations shown in Fig. 5 demonstrate that the signal ratio can lead the warnings ahead of time compared to the referenced methods, which further warrants the feasibility of adding signal ratio to current warning systems.

## V. CONCLUSIONS

A disease outbreak can affect a large population and spread over multiple countries or continents in a short period, posing significant health threats to humanity [10]. Early warning is critical for government agencies to contain the disease outbreak. The extensive application of social media opens up new insights to promote early warning in many scenarios, and understanding the online information created from particular times and places can provide valuable resources that can be incorporated into early warning systems. Previous studies have demonstrated the potential of using social media data to signal early warning by analyzing symptoms-related postings or tracking online search queries. While those attempts based on classifying symptoms-related postings can provide real-time surveillance, they may be limited to the situation where the symptoms are widely reported in the communities. While tracking online search queries can predict the risks ahead of the outbreak, they may provide limited insights when the aberrations are not discernible at the beginning of a crisis. Further, with our reviewed studies, there has been limited discussion on how to quantitatively analyze public awareness through social media data to enhance early warning.

To explore the potential of social media data, this study conducts a retrospective analysis of the U.S. COVID-19 pandemic and investigates more than 14 million related tweets from January 20 to March 10, 2020. With the aid of natural language processing and machine learning techniques, this study classifies each related tweet into either a signal or a non-signal and further proposed a parameter “signal ratio” to detect the warning signs in the early stages. Following are the highlighted findings. The signal ratio presented an aberrational pattern of “continuous warning signs” since February 20, which unfolded the crisis much earlier than the official statement. In particular, the signal ratio exhibited a peak value on February 25 (about 0.25), indicating that a large number of Twitter users had already been aware of the hazards. Even from late January to early February, when the topic was not widely discussed online, the exposure to other countries’ news on social media enabled a proportion of users to realize the COVID-19 crisis and its potential threats to the U.S. public. Moreover, the emerged pattern on Twitter shifted from concerns about external situation (e.g., virus mechanism, other countries’ outbreaks) in early February to an extremely pessimistic mood about internal situation (e.g., lack of testing kits and equipment, underestimated risks) in early March. Last, through the validation with other referenced methods based on media news, this study demonstrated that the signal ratio could detect the outbreak hazards 16 days earlier.

The contribution of this study mainly lies in two aspects. From a methodological perspective, this study presents a quantification process of dealing with social media data and offers an approach to quantitively estimate the awareness based on the classifications of signal and non-signal tweets. Unlike the methods based on frequency count (e.g., tracking search queries, identifying behavior changes based on word patterns), this study applies machine learning methods to identify the signals from textual contents on social media. Thus, the signal ratio reflects people’s awareness of the crisis and can detect warning signs even with low discussion volume in the early stages.

From a practical perspective, this study demonstrates the potential of aggregating public opinions via social media to signal early warning. Some useful information on social media can serve as the whistles for the disease outbreak in a broader sense. A typical case is that, before the COVID-19 outbreak, a Chinese doctor shared the information of a suspected respiratory syndrome on social media, which raised early awareness about COVID-19 infections [16]. This study also illustrates a case where public opinions are of importance for government agencies to take early measures when facing an unprecedented disease outbreak. Overall, this approach has the advantages of rapidity, quantity, spatial converge, and certain foresight in unfolding the crisis, and it can feed into current warning systems that can help government agencies enhance early warning in a timely manner.

Limitations to this study need to be addressed in future applications. First, this signal ratio may not be applicable for long-term use, especially when a place has witnessed the outbreak. This is because people’s discussion on social media may switch to other aspects of the pandemic, and the topic regarding the risks (i.e., warning signals) of the pandemic may not be actively discussed. Second, this study sets 0.1 as the critical value of the signal ratio (i.e., the fraction of “signal” tweet volume) for a warning sign. As there is no firm reference to support this setting in the social media context, the credibility of the warning level calibration may require discussions in a more general case. Third, given that this proposed signal ratio is discussed in the COVID-19 context, the training dataset may need to be adjusted when the model is applied to signal warnings for other pandemics. Last, the TF-IDF technique may not be suitable for the situation where the tweet data are very different from the training tweet data since this method does not take the semantics or positions of words into account.

Ongoing and future work will assess other text classification models by using advanced techniques (e.g., deep learning classifiers, word embedding methods) to build the pipelines. Other potential study direction will focus on evaluating the tipping point regarding public awareness and social media context. From the angle of application, we will collect tweets relative to other types of pandemics or epidemics (e.g., measles, swine influenza) and apply the model to detect early warnings in other health crises. This proposed model can supplement current early warning systems and further aid the public, the healthcare professionals, and government agencies to prepare for future health crises.

## Data Availability

The data can be obtained upon reasonable request from authors.

## DATA AVAILABILITY

The data that support the findings of this study are retrieved from the Twitter Inc., but restrictions apply to the availability of these data, which were used under license for the current study, and so are not publicly available. The tweet IDs are however available from the authors upon reasonable request, but the original tweets are available with permission of the Twitter Inc. The data that support the findings of this study are also available from the corresponding author upon reasonable request.

